# Towards safer risperidone prescribing in Alzheimer’s disease

**DOI:** 10.1101/2020.08.12.20173310

**Authors:** Suzanne Reeves, Julie Bertrand, Hiroyuki Uchida, Kazunari Yoshida, Yohei Otani, Mikail Ozer, Kathy Y Liu, Elvira Bramon, Robert Bies, Bruce Pollock, Robert Howard

**Author notes:** Correspondence to: Suzanne Reeves, Professor of Old Age Psychiatry and Psychopharmacology, Division of Psychiatry, Faculty of Brain Sciences, University College London, 6th Floor, Maple House, 149 Tottenham Court Road, London W1T 7NF.

## Abstract

**Background:** In the treatment of psychosis, agitation and aggression in Alzheimer’s disease (AD), guidelines emphasise the need to ‘use the lowest possible dose’ of antipsychotic drugs, but provide no information on optimal dosing.

**Aims:** This analysis investigated the pharmacokinetic profiles of risperidone and active metabolite, 9-hydroxy (OH)-risperidone, and how this related to emergent extrapyramidal side effects (EPS), using data from The Clinical Antipsychotic Trials of Intervention Effectiveness-AD study.

**Method:** A statistical model, which described the concentration-time course of risperidone and 9-OH-risperidone, was used to predict peak, trough and average concentrations of risperidone, 9-OH-risperidone and ‘active moiety’ (combined concentrations) (108 CATIE-AD participants). Logistic regression was used to investigate the associations of pharmacokinetic biomarkers with EPS. Model based predictions were used to simulate the dose adjustments needed to avoid EPS.

**Results:** The model showed an age-related reduction in risperidone clearance (p<0.0001), and estimated that 22% of patients had slower active moiety clearance (concentration-to-dose ratio 20.2±7.2 versus 7.6±4.9 ng/mL per mg/day, Mann Whitney U, *p*<0.0001). Higher average and trough 9-OH-risperidone concentrations (*p*<0.0001), and lower Mini-Mental State Examination (MMSE) scores (p<0.0001), were associated with EPS. Model based predictions suggest the optimum dose ranged from 0.25mg/day in those aged 85 years with MMSEs of 5, to 1mg/day in those aged 75 years with MMSEs of 15, with alternate day dosing required for those with slower drug clearance.

**Conclusions:** Our findings argue for age- and MMSE -related dose adjustments and suggest that a single plasma sample could be used to identify those with slower drug clearance.

## Introduction

### Antipsychotic drug use in Alzheimer’s disease

Alzheimer’s disease affects around 35 million people worldwide, fifty percent of whom will experience psychosis symptoms (delusions and hallucinations). (1) Psychosis symptoms are often distressing, increase the risk of aggression towards caregivers, predict faster cognitive and functional decline, and reduce ability to live independently (2). Although symptoms sometimes respond to psychosocial interventions, for those with severe persistent symptoms, antipsychotic medication is required to reduce distress and associated risks. (3) The best evidence of efficacy is for second generation antipsychotic drugs. (4) However, concerns about side-effects (sedation, falls, parkinsonism, and stroke) and increased mortality in people with dementia, (5, 6) particularly in those aged over 80 years, (7) has led to a restriction in prescribing. In England, National Institute for Clinical Excellence (NICE) guidance emphasises the need to treat with ‘the lowest effective dose for the shortest possible time’ but provides little practical information on the optimal dose range for individual drugs. We have shown that amisulpride therapeutic plasma concentrations for the treatment of AD psychosis (40-100 ng/mL), are lower than those recommended for the treatment of schizophrenia (100-320 ng/mL), due to a leftwards shift in the dopamine D_2/3_ receptor concentration-occupancy curve. (8) These findings raise questions regarding the mechanisms of antipsychotic sensitivity in AD and suggest that, for amisulpride at least, 50 mg/day (compared to 400-800mg/day in young adults), may optimally balance the risks and benefits of treatment. (9) It is, however, not clear how far we can extrapolate this approach to other antipsychotic drugs.

### Pharmacokinetics and consensus guidance on risperidone prescribing

Risperidone, an antipsychotic drug with high affinity for dopamine D_2/3_ and serotonin 5HT_2A_ receptors, is the only drug licensed for short-term use in the treatment of aggression and psychosis in dementia in the European Union, and is typically prescribed across a 0.5-2 mg/day dose range in this indication. (5, 10) Oral risperidone has high (70-85%) bioavailability and is extensively metabolized in the liver by cytochrome P450 2D6 (CYP2D6) to the active metabolite 9-hydroxy (OH)-risperidone. (11) Peak concentrations of risperidone and 9-OH-risperidone are reached after 1 and 3 hours respectively, and steady state concentrations of the active moiety (combined concentrations of risperidone and 9-OH-risperidone) are achieved after 4-5 days. The elimination half-life (t_1/2_) of risperidone is dependent on multiple factors. Genetic variation in CYP2D6 genotype accounts for around 50% of the variability in risperidone concentrations (t_1/2_ for risperidone ranges from 4.7 hours in normal metabolisers to 22 hours in poor metabolisers); with age, hepatobiliary dysfunction, and use of CYP2D6 inhibitors (paroxetine) or inducers (carbamazepine) further contributing to variability (12, 13). The metabolite is renally excreted, with a t_1/2_ of 20 hours; increased to 25 hours in the over-65s and in moderate renal failure.

Consensus guidelines, based on therapeutic drug monitoring, (11, 14) pharmacokinetic modelling, (15) and imaging of striatal D_2/3_ receptor occupancy (16) in risperidone treated patients with schizophrenia, recommend active moiety concentrations of 20–40 ng/mL (3-6 mg/day), (15) as higher concentrations increase occupancy beyond 80% and increase the risk of extrapyramidal side-effects (EPS). Recent guidance on personalised risperidone prescribing advocates dose reductions for those with slower clearance of the active moiety, indexed by concentration to dose (C/D) ratios of the active moiety over 14 ng/mL per mg/day. (17) There is a lack of empirical data from people with AD.

### Aims

This analysis aimed to combine pharmacokinetic and clinical outcome data from The Clinical Trials of Intervention Effectiveness in Alzheimer’s disease (CATIE-AD) study, (18) with the following objectives:

1. To investigate sources of variability in plasma concentration-time profiles of risperidone and 9-OH-risperidone, using an approach that allowed estimation of risperidone clearance (metabolism) in distinct subpopulations.
2. To estimate pharmacokinetic indices (peak, trough and average concentrations of risperidone, 9-OH-risperidone and active moiety) for each individual, across the prescribed dose range.
3. To investigate the relationship between the above pharmacokinetic indices with EPS.

## Method

### Data source

CATIE-AD (6, 18) was a randomized, double-blind, parallel group study comparing olanzapine, quetiapine, risperidone and placebo in the treatment of psychosis and aggression in AD (Clinicaltrials.gov identifier: NCT00015548). In phase 1, participants were randomized to receive risperidone, olanzapine, quetiapine, or placebo in a 1:1:1:1 allocation ratio, with study physicians having a choice of two capsule strengths (low or high). For risperidone, this corresponded to 0.5 mg or 1.0 mg. Doses were adjusted as clinically indicated by study physicians. If the physician judged that the patient’s response was not adequate at any time after the first 2 weeks, then treatment could be discontinued, with a further decision point being made at 12 weeks. Patients with an adequate response continued treatment for up to 36 weeks. Patients whose initial treatment was discontinued during phase 1 could be enrolled in phase 2 and randomly assigned under double-blind conditions to receive one of the antipsychotic drugs to which they were not initially assigned, or to receive citalopram. In phase 3, treatment was prescribed in an open manner. Within each phase, plasma drug concentration was measured at 2, 4 and 12 weeks, or when a medication switch was made (6, 18).Clinical assessment (Baseline, every 2-4 weeks during dose titration) included the Simpson Angus Scale (SAS), (19) and Barnes Akathisia Scale (BAS). (20) Plasma concentrations of risperidone and the active metabolite 9-OH-risperidone were determined using a liquid chromatography-tandem mass spectrometry method with a detection limit of 0.1 ng/mL. The authors assert that all procedures contributing to this work comply with the ethical standards of the relevant national and institutional committees on human experimentation and with the Helsinki Declaration of 1975, as revised in 2008. Written informed consent was obtained from all patients. (6) A comprehensive plan was developed to ensure that all institutional, National Institute for Health, and federal regulations concerning informed consent were fulfilled. The plan included careful assessment of risks and benefits, review by the CATIE protocol and ethics committees, and re-view by the National Institute of Mental Health Data Safety and Monitoring Board. (18)

### Data Extraction

Data available from risperidone treated participants included study identification number, phase, visit, dose (mg), timing of blood draw (hours post dose), number of days of treatment, dosage interval (daily), physiological characteristics (age, gender, height, weight, ethnicity (coded as white/other), and smoking (currently smoking or not), Mini-Mental State Examination (MMSE) scores, and plasma concentrations of risperidone and 9-OH-risperidone (ng/mL). Treatment emergent EPS were coded as present if SAS total scores were six or more, or BAS global scores were two or more at follow-up, in individuals with Baseline SAS ratings less than six and BAS scores less than two. Only participants without Baseline EPS were included in our analysis of outcome data. Information was cross-checked with adverse event data, to establish if the event was reported during the relevant phase of treatment; coded under Medical Dictionary for Regulatory Activities (MedDRA 4.0) (preferred terms ‘Parkinsonism’, ‘Parkinsonism aggravated’, ‘cogwheel rigidity’, ‘masked facies’, ‘bradykinesia’, ‘tremor’, ‘extrapy rami dal disorder’, ‘drooling’, ‘gait festinating’, akinesia, akathisia, or tardive dyskinesia); and rated as possibly or probably related to treatment. Data extracted on other adverse events included sedation (preferred terms ‘sedation’, ‘somnolence’, ‘hypersomnia’, ‘lethargy’), falls (preferred term ‘fall’), and postural hypotension (preferred term ‘hypotension or hypotension not otherwise specified’). Rating scales and recorded adverse events were checked for consistency with pharmacokinetic data, using phase, number of days treatment, and the timing of blood sampling.

### Statistical Analysis

#### Demographics

Demographic data were analysed using statistical package for social sciences version 22.0. Mann Whitney U tests were used to describe group comparisons. Chi-squared tests were used to compare frequencies between groups.

#### Pharmacokinetic model development

Plasma concentration-time profiles of risperidone and 9-OH-risperidone were evaluated using a statistical model (21) that linked parent risperidone and metabolite 9-OH-risperidone via a metabolism rate constant (*km*), with the following parameters: i) Risperidone clearance (*CL_RISP_*), ii) Risperidone volume of distribution (*V_RISP_*), iii) Absorption rate constant *(ka)*, iv) 9-OH risperidone volume of distribution (*V_9-OH-RISP_*), and v) 9-OH-risperidone clearance (*CL_9-OH-RISP_*). The analysis estimated fixed effects (parameters describing dose-concentration relationships), and random effects, comprised of inter-individual variability (difference between individual and predicted model parameter values for the sample), and residual variability (system noise, dosage history errors). (22) Model development was carried out using Monolix software (version 2018r; www.lixoft.eu). Parameters were estimated using an iterative approach which provided maximum likelihood estimates and standard errors. Plasma concentration was converted from ng/mL to mcg/L for use in model building. The model allowed estimation of the probability of there being more than one subpopulation in relation to risperidone clearance, by including a latent covariate. No assumption was made that latent categories corresponded solely to CYP2D6 genotype, as multiple factors contribute to hepatic metabolism in older people. Residual variability was estimated separately for risperidone and 9-OH-risperidone. Covariates (height, age, gender, smoking, ethnicity, weight) were incorporated in a stepwise manner, through visual inspection of covariate plots and regression analysis in R for categorical covariates. Models were evaluated using goodness-of-fit criteria, including diagnostic scatter plots, visual predictive checks, degree of shrinkage, change in inter-individual variability, model precision, and approximate likelihood ratio tests. A change in log likelihood estimate was considered significant if =>4 (equivalent to *p*<0.05, one degree of freedom), and accompanied by no change or a decrease in Bayesian Information Criteria.

#### Pharmacokinetic biomarkers and clinical outcome

Model based estimates were used to calculate peak, trough, and average concentrations of risperidone, 9-OH-risperidone and active moiety (their combined concentrations) for each individual, across the dosage interval. Concentration-to-dose ratio for the active moiety was calculated from trough estimates, to allow comparison with recommendations regarding personalised dosing of risperidone. (17) Each pharmacokinetic biomarker was individually considered as an independent variable (regressor) in a binary logistic model which described the probability of EPS.. The model accounted for random effects, and adjusted for potential confounders (age, sex, MMSE, height, weight). Best fit models were used to simulate and predict plasma concentrations and probability of response and EPS, accounting for factors that contributed to variability.

## Results

### Sample characteristics

Of 110 risperidone treated patients, 65 (59.1%) were randomised to risperidone treatment in phase one, 31 (28.2%) in phase two, and 14 (12.7%) in phase three (188 plasma samples, collected 26.9±69.9 hours post dose). After excluding four samples, taken after 180 hours (above six half-lives post dose), data from 108 patients remained (52 (47.3%) men, aged 78.4±6.7 years, weight 68.9±14.7 kg, height 1.6±0.1m, MMSE 14.6 ±6.2); sampled 18.1±26.8 hours post dose, after 92.4±76.8 days treatment with 1.0±0.7 mg/day of risperidone (risperidone plasma concentrations 2.4±3.1ng/mL, 9-OH-risperidone plasma concentrations 10.0±8.4 ng/mL).

Eight participants with Baseline SAS scores of six or more (indicating EPS prior to commencing risperidone), were excluded from the analysis of outcome data. Those with Baseline EPS had greater global cognitive impairment (MMSE 7.8±7.0 versus 15.2±5.8, Mann Whitney U, *p*<0.0001) but there were no differences in other characteristics (Table 1). Treatment emergent EPS occurred in 14 (14%), sedation in 13 (13%), falls in five (5%), postural hypotension in two (2%) and ECG abnormalities in three (3%) patients (Table 1). Those with EPS were prescribed a higher risperidone dose (1.7±0.9mg versus 0.9±0.5mg; Mann Whitney U, *p*<0.003), had lower MMSE scores (10.2±4.2 versus 16.0±5.6, Mann Whitney U, *p*<0.0001) and a greater proportion (5 (37.5%) versus 6 (7.0%) patients) were treated with concomitant anti-depressant medication (trazodone) (chi-squared *p*=0.007, odds ratio = 7.4, 95% CI 1.9-29.2).

**Table 1.**
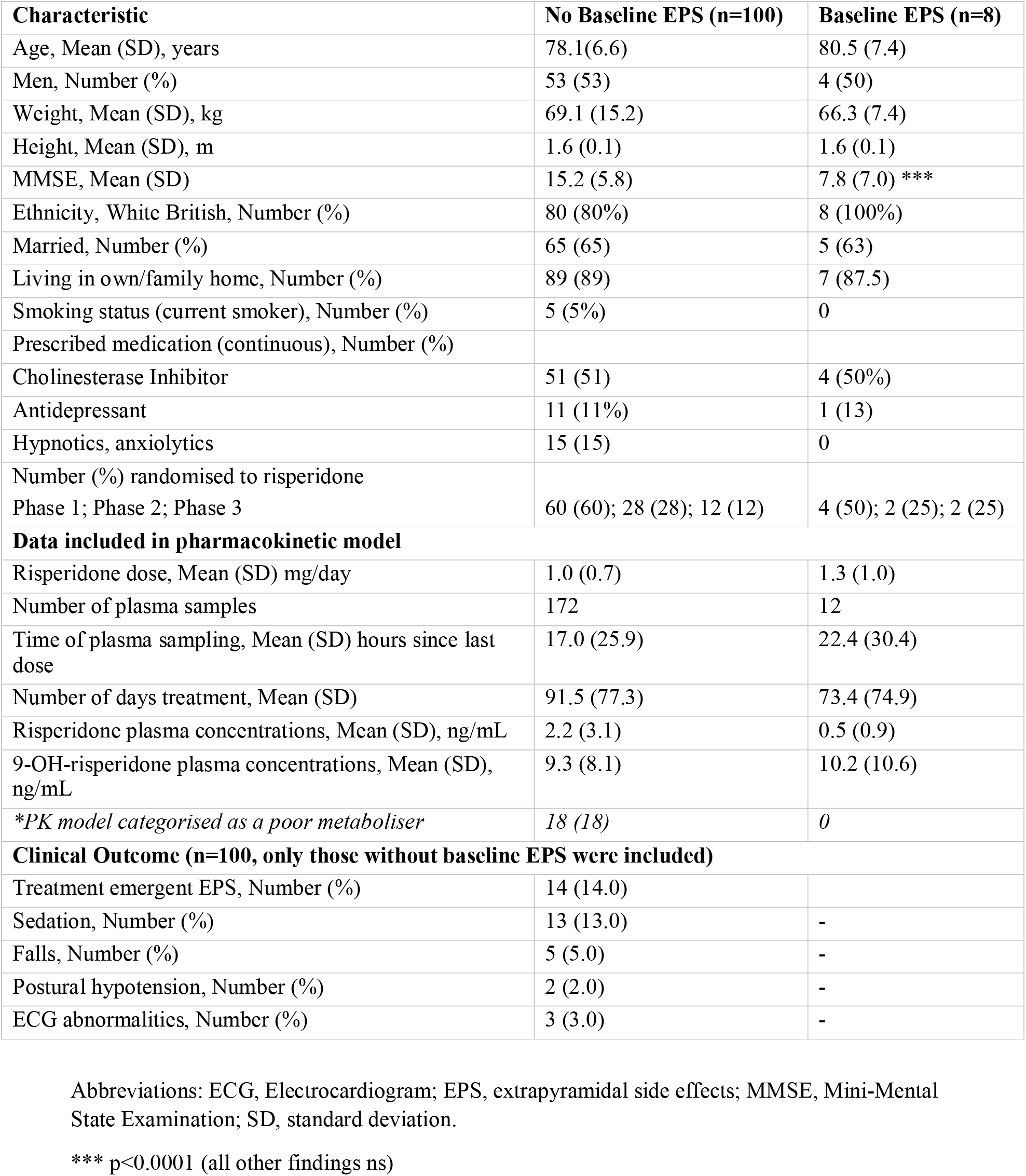
Demographic and Clinical Characteristics of Risperidone-Treated CATIE-AD Participants.

### Pharmacokinetic model

The base model included a latent covariate with two categories (the model failed to converge using a covariate with three categories). Parameters were estimated with good precision, apart from V _9-OH-RISP_ (relative standard error 60.1%). Residual variability was 0.1 mcg/L (56.2%) for risperidone and 0.7 mcg/L (28.2%) for 9-OH-risperidone. Covariate testing identified a significant contribution of log transformed age (tAge) to the variability in risperidone clearance (β =-0.3, *p*=9.13e-04). Inclusion of an age effect on *CL_RISP_* increased the precision of the model (Supplementary Table 1) and reduced the estimated probability of being in latent category 1 from 32% to 22%. Stepwise testing of other covariates on clearance parameters did not improve the precision or model fit.

Model based predictions, based on the mean age (78.4 years) of the sample, suggested that for patients assigned to latent category one, risperidone clearance was 8.7L/hr (t_1/2_ 22 hours), compared to 34.2L/hr (t_1/2_ 5 hours) for those in latent category two. Patients in latent category one were thus considered to represent ‘functionally poor’ metabolisers (PM). Predictions based on the observed contribution of age to risperidone clearance estimated that, for those aged 88 years, risperidone clearance would be reduced by 22% (t_1/2_ 28.8 hours in PM and 6.5 hours in functionally normal metabolisers (NM). Based on V _9-OH-RISP_ and *CL_9-OH-RISP_*, t_1/2_ 9-OH-risperidone was 27 hours. Visual predictive checks, (VPCs) shown as percentile plots, superimposed on observed data, are shown in Supplementary Fig 1.

### Pharmacokinetic biomarkers and functional metaboliser status

Of the 100 participants included in the analysis of clinical outcome, 18 were categorised as PM. There were no differences in clinical or demographic or clinical variables in PM and NM. Cholinesterase inhibitors were prescribed in a higher proportion of PM (four (77.8%) versus 37 (45.1%), chi squared *p*=0.01, odds ratio 1.27, 95% confidence interval (CI) 1.04-1.53), and a higher proportion of PM were women (14 (77.8%) versus 39 (47.6%), chi squared *p*= 0.02, odds ratio 1.24, 95% CI 1.04-1.49). PM were prescribed a lower dose of risperidone (0.7±0.4 mg/day versus 1.1±0.7mg/day, Mann Whitney U, *p*=0.007), and had higher concentrations across all pharmacokinetic biomarkers (Supplementary Table 2). Trough active moiety concentration-to-dose ratio was markedly increased in PM (20.2±7.2 versus 7.6±4.9 ng/mL per mg/day, Mann Whitney U, *p*<0.0001), shown in Fig 1. There were no differences in the proportion of participants with emergent EPS in PM and NM.

**Fig 1.**
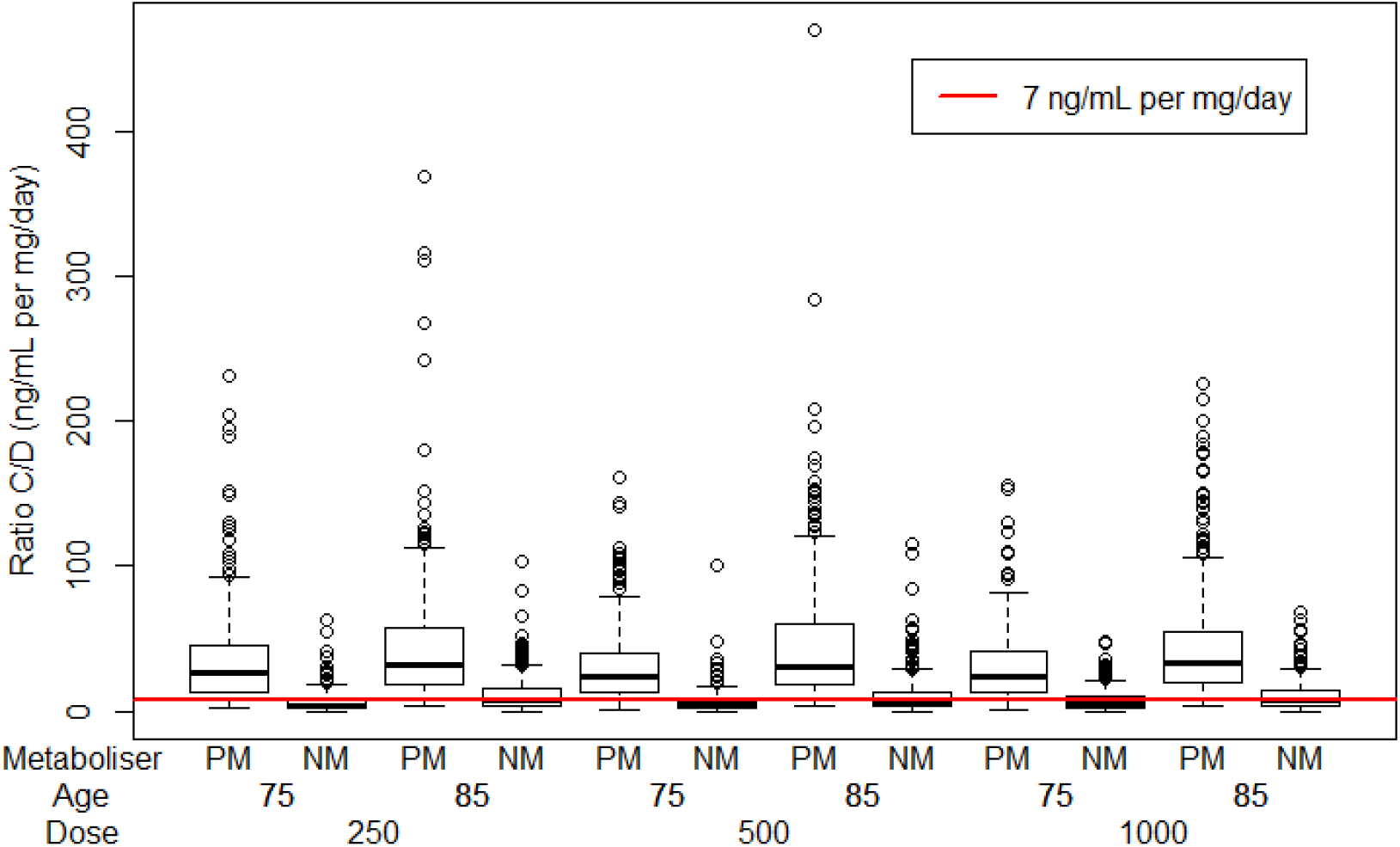
Active moiety concentration to dose ratio. Estimated concentration to dose (C/D) ratios for the active moiety (ng/mL per mg/day) are shown at trough in those categorised as functionally poor (PM) and functionally normal (NM) metabolisers, aged 75 and 85 years, prescribed 250, 500 and 1000mcg risperidone daily. The red line (7ng/mL per mg/day) represents typical estimates for C/D in a reference group, based on therapeutic drug monitoring studies of risperidone.

### Pharmacokinetic biomarkers and clinical outcome

Associations between individual pharmacokinetic biomarkers and EPS are detailed in Table 2. Pharmacokinetic biomarkers showed a significant association with EPS, achieving greatest significance in relation to trough and average concentrations of 9-OH-risperidone and the active moiety (Wald test *p* <0.0001). MMSE and height also contributed to the best fit regression models (*p*<0.0001).

**Table 2.** Pharmacokinetic Biomarkers and Emergent EPS (n=100)

| **Table 2. Pharmacokinetic Biomarkers and Emergent EPS (n=100)** |  |  |
| --- | --- | --- |
| **Regressor #** | **Wald test p value** | **BIC** |
| **Peak plasma concentrations, ng/mL** |  |  |
| Risperidone | 0.04 | 73.0 |
| MMSE | ns |  |
| 9OH-Risperidone | <0.0001 | 65.9 |
| MMSE | <0.0001 |  |
| Active Moiety | 0.03 | 66.6 |
| MMSE | ns |  |
| **Trough plasma concentrations, ng/mL** |  |  |
| Risperidone | ns | 74.8 |
| MMSE | ns |  |
| 9OH-Risperidone | <0.0001 | 60.6 |
| MMSE | <0.0001 |  |
| Active Moiety | <0.0001 | 61.7 |
| MMSE | <0.0001 |  |
| **Average plasma concentrations, ng/mL** |  |  |
| Risperidone | ns | 73.3 |
| MMSE | 0.03 |  |
| 9OH-Risperidone | <0.0001 | 60.6 |
| MMSE | <0.0001 |  |
| Active Moiety | <0.0001 | 61.9 |
| MMSE | <0.0001 |  |

Binary logistic regression models accounted for random effects, and adjusted for potential confounders including gender and log transformed age, MMSE, height, and weight.
A backward method was used, which removed variables that did not contribute to the model (significance threshold $p < 0.05$ ). For each best fit model, the $\beta$ regressor effect coefficient (standard error) value was used to calculate a Wald statistic, and its $p$ value, based on the chi squared statistic.

Model-based simulations (see Fig 2) suggest that for NM aged 75 years with an MMSE of 15, 1mg/day risperidone would be associated with minimal risk of EPS. For those aged 75 years with an MMSE of 5, a dose reduction to 0.5mg/day would be required. For those aged 85 years, the dose would need to be reduced by 50% to achieve equivalent plasma concentrations, and PM would require very low alternate daily dosing (0.25-0.5mg/48 hours in those aged 75 years, and 0.125-0.25mg/48 hours in those aged 85 years).

For completeness, the same analysis was carried out in relation to sedation, but there were no significant associations. The number of participants with other emergent side effects was too small to investigate through the use of logistic regression.

**Fig 2.**
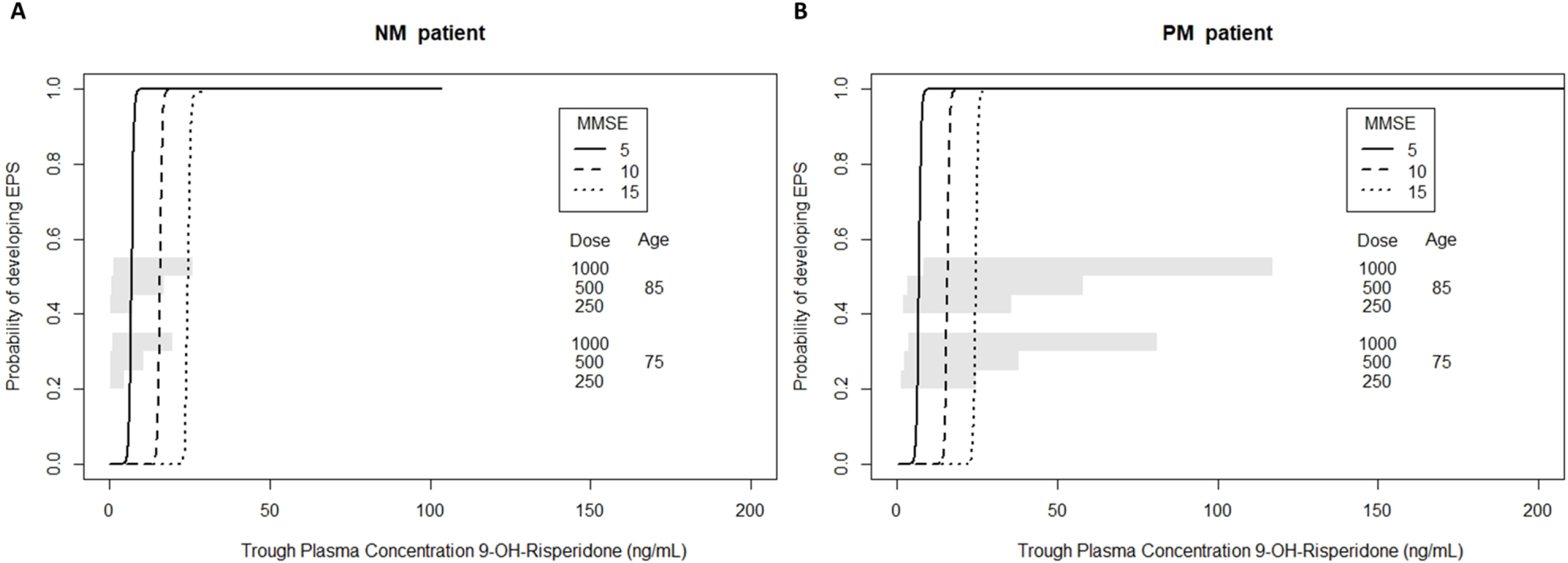
Simulated trough 9-OH-risperidone concentrations and EPS. Simulated trough 9-OH-risperidone concentrations and the probability of extrapyramidal side effects (EPS) are shown for a population of 100 people in each of the following categories: 75 or 85 years old; with an MMSE score of 5, 10 or 15; prescribed 250, 500 or 100mcg risperidone daily in A) Functionally normal metabolisers (NM) and B) Functionally poor metabolisers (PM).

## Discussion

### Age and functional metaboliser status contribute to pharmacokinetic variability

The consensus, based on meta-analyses of placebo controlled trials of risperidone in people with psychosis in AD, is that 1 mg/day risperidone may optimally balance efficacy and adverse effects. (4-6, 10) However, meta-analyses can only inform the ‘average’ dose requirements, but are less able to identify subgroups who are most susceptible to side-effects. In this analysis we have investigated the pharmacokinetic (dose-concentrations) and pharmacodynamic (concentration-outcome) contribution to EPS, to guide safer prescribing. We estimated that 22% of patients were functionally poor metabolisers and observed an independent effect of age on risperidone clearance.

Pharmacokinetic biomarkers were robust predictors of EPS, with higher trough and average concentrations of 9-OH-risperidone providing the best fit for the data. Lower MMSE, a marker of more severe global cognitive impairment, was an independent predictor of EPS. Model based simulations suggest that, for NM aged 75 years with an MMSE of 15 (moderate AD severity), 1mg/day would be associated with minimal risk of EPS, but a dose reduction to 0.5mg/day would be required for those with an MMSE of 5. For those aged 85 years, a 50% dose reduction would be required and, for PM, alternate day dosing would be required to avoid excessive exposure.

Drug metabolism is a major contributor to pharmacokinetic variability (23) and, in older adults, the relative importance of genotype is difficult to disentangle from physiological and clinical factors. (17) In the absence of information on *CYP2D6* genotype, the extent of any genetic contribution to functional metaboliser status is unclear, although estimates of risperidone clearance in PM and NM were broadly consistent with estimates for *CYP2D6-*predicted ‘poor’ and ‘extensive’ metabolisers. (13) Age was independently associated with risperidone clearance and, when incorporated into the model, led to the reassignment of six patients who were initially categorised as PM. Although previous research has not shown an effect of age specifically on risperidone clearance, (13) a 30% decrease in hepatic metabolism in those aged over 70 years has been observed for other CYP2D6 substrates (24) and it is thus likely that our findings are explained by the older age of CATIE-AD participants. Similarly the absence of an association between age and 9-OH-risperidone clearance reflects the older age of CATIE-AD participants compared to previous analyses.

### Mechanistic insights into emergent EPS

Statistical modelling of the relationship between risperidone active moiety plasma concentrations and D_2/3_ receptor occupancy (25) in adults with schizophrenia, suggests that trough concentrations of 10.5-38.2 ng/mL are associated with 60-78% occupancy in the striatum, and 6.5 ng/mL (95% CI, 3-10 ng/mL) is associated with 50% occupancy. In CATIE-AD participants, EPS emerged from trough active moiety concentrations of 3.4 ng/mL (of which 3.2 ng/mL was 9-OH-risperidone), and concentrations exceeded 10ng/mL (60% occupancy) in eight of 14 patients with EPS. In the absence of occupancy data, it is unclear whether the emergence of EPS at such low concentrations signifies a leftwards shift in the concentration-occupancy curve, similar to that observed during amisulpride treatment. (8) This is possible, as risperidone and 9-OH-risperidone are substrates for P-glycoprotein, (26) a blood brain barrier efflux transporter that is marked reduced older people with AD. (27) However age or disease-specific changes in brain drug distribution, clearance, and competition with endogenous dopamine at receptor sites need to be considered. (8)

Pharmacodynamic changes (reduced D_2/3_ receptor reserve, altered signal transduction) which lead to a greater functional outcome for a given occupancy are also important. We have previously observed EPS at low striatal D_2/3_ receptor occupancies (60% compared to 80% in young people), in risperidone-treated older people with schizophrenia, (28, 29) and amisulpride-treated older people with psychosis in AD. (8) Given the error margin of occupancy predictions, we cannot rule out the possibility that occupancy was under-estimated in a proportion of those with active moiety concentrations less than 10ng/mL. (25)

To our knowledge, this is the first demonstration of the contribution of MMSE score, a marker of increasing dementia severity, to the risk of antipsychotic related EPS. An association between agitation, antipsychotic use and death in those with more severe dementia has however been reported in a recently published cohort study (30). The mechanisms of antipsychotic-induced EPS are not fully understood, but D_2/3_ receptor antagonism of inhibitory dopaminergic inputs to striatal medium spiny neurones and cholinergic interneurones may play a key role. (3) It is unclear whether the risk of EPS in those with lower MMSE scores reflects greater in networks that modulate motor control, or is associated with as yet unidentified factors that potentiate EPS.

### Limitations

Limitations to the analysis include sparse sampling, which meant it was not possible to estimate within-subject variability in clearance of risperidone or 9-OH-risperidone, or to investigate the contribution of concomitant medications to pharmacokinetic variability. Neither was it possible to investigate the contribution of comorbid medical conditions to variability in pharmacokinetics or emergent side effects. We cannot account for the fact that those with emergent EPS were more likely to have been prescribed concomitant trazodone, given the small sample size and uncertain exposure (dose, continuity of the prescribed drug) of individual participants to trazodone. However, we cannot rule out the possibility of potential drug-drug interactions, including an interaction with P-glycoprotein (trazodone is a substrate), or potentiated EPS through the inhibition of serotonin uptake. (3) Other limitations relate to the CATIE-AD study design, which may have reduced our ability to detect associations between pharmacokinetic indices and clinical outcome. This includes flexibility in starting dose (low or high), the option of making adjustments or of discontinuing a phase, based on clinician judgement.

### Dose adjustments needed to avoid treatment emergent EPS

This analysis represents a step towards safer risperidone prescribing and argues strongly for age- and MMSE-related dose reductions. From a pragmatic perspective, clinicians should ‘start low go slow’ (0.5-1mg/day) in those aged 75 years with moderate stage AD, and ‘start low, stay low’ (maximum 0.5mg/day) in those aged 75 years with severe AD. For those aged 85 years, the dose should be halved, and alternate daily dosing considered if side effects emerge, as it is likely that the person has slower active moiety clearance.

Personalised prescribing should ideally incorporate knowledge of genetic, environmental and personal variables to determine dosing. This is currently happening in clinical practice and there has been a lack of empirical data in older people to justify the use of routine therapeutic drug monitoring. Consistent with a recently proposed personalised prescribing algorithm, CATIE-AD participants categorised as PM had high active moiety concentration-to-dose ratios. (17) Therapeutic drug monitoring thus offers the opportunity to guide dose adjustments with greater precision, as this measure could be derived from a single plasma sample, to identify those with slower drug clearance. Future research should test our proposed dose predictions and could evaluate the clinical utility of therapeutic drug ‘screening’ (measurement from a single plasma sample).

## Data Availability

Data used in the preparation of this article were obtained from the limited access datasets distributed from the NIH supported Clinical Antipsychotic Trials of Intervention Effectiveness in AD (CATIE-AD) (MH64173, MH90001). This is a multisite, clinical trial of people with psychosis or agitation in the content of AD, which compared the effectiveness of randomly assigned medication treatment. This manuscript reflects the views of the authors and may not reflect the opinions or views of the CATIE Study Investigators or the NIH. No funding was provided for this analysis. Suzanne Reeves had full access to the data included in this analysis, and had final responsibility for the decision to submit for publication.

## Acknowledgements

Data used in the preparation of this article were obtained from the limited access datasets distributed from the NIH supported “Clinical Antipsychotic Trials of Intervention Effectiveness in AD” (CATIE-AD) (MH64173, MH90001). This is a multisite, clinical trial of people with psychosis and/or agitation in the content of AD, which compared the effectiveness of randomly assigned medication treatment. This manuscript reflects the views of the authors and may not reflect the opinions or views of the CATIE Study Investigators or the NIH. No funding was provided for this analysis. Suzanne Reeves had full access to the data included in this analysis, and had final responsibility for the decision to submit for publication.

## Contributors

SR led the study design, carried out data extraction, carried out data analysis and led on the writing of the paper; JB, HU, RB, BP and RH gave input into the study design and analysis plan; KY, YO and KL extracted data on clinical outcome measures; MO extracted data on adverse events; JB supervised SR in the analysis of the data, carried out simulations, and led on data presentation; EB and RH gave input into the interpretation and presentation of pharmacokinetic data; all authors contributed to the interpretation of the clinical findings and the writing of the paper, and approved the submitted manuscript.

## Competing interests

There are no competing interests

## Funding

R.H. is supported by the NIHR University College London Hospital Biomedical Research Centre (UCLH BRC)

## Supplementary Material

**Supplementary Table 1.** Pharmacokinetic Model for Risperidone and 9-hydroxy-risperidone (n=108)

|  |  |  |  |  |
| --- | --- | --- | --- | --- |
| Parameters | Base Model |  | Final Model |  |
| Fixed Effects | Parameter estimates | RSE (%) | Parameter estimates | RSE (%) |
| Ka (/hr) (fixed) | 2 | - | 2 | - |
| V<sub>RISP</sub> (L) | 281 | 16 | 272 | 16.4 |
| CL<sub>RISP</sub> (L/hr) | 11.1 | 26.3 | 8.87 | 24.8 |
| $\beta$ Latent category (2) CLp | 1.3 *** | 20.2 | 1.4 *** | 15.8 |
| $\beta$ tAge CLp | - | - | -3.1 *** | 30.2 |
| V<sub>9-OH-RISP</sub> (L) | 1703 | 60.1 | 2030 | 49.2 |
| CL<sub>9-OH-RISP</sub> (L/hr) | 53.2 | 13.6 | 52 | 13.5 |
| Interindividual variability, |  |  |  |  |
| $\omega$ V<sub>RISP</sub> % | 42.5 | 32.7 | 45.2 | 39.2 |
| $\omega$ CL<sub>RISP</sub> % | 53.8 | 23.5 | 46.8 | 20.9 |
| $\omega$ V<sub>9-OH-RISP</sub> % | 133 | 40.2 | 97.0 | 47.5 |
| $\omega$ CL<sub>9-OH-RISP</sub> % | 55.4 | 16.4 | 55.6 | 17.6 |
| Residual unexplained variability |  |  |  |  |
| $\sigma$ Risperidone mcg /L (%) | 0.1 (56.2) | 36.6 (15.3) | 0.1(55.6) | 21.1 (11.6) |
| $\sigma$ 9-OH-risperidone mcg/L (%) | 0.7 (28.2) | 35.4 (16.3) | 0.7 (28.7) | 39.0 (15.0) |
| Probability of being in latent category, % | 31.6 | 35.4 | 21.7 | 40.2 |
| -2 x Log likelihood/ Bayesian Information Criteria | 1700/1766 |  | 1688/1759 |  |

Abbreviations: *Ka* -first-order absorption rate constant (fixed at 2); V<sub>RISP</sub> - Volume of distribution for risperidone; CL<sub>RISP</sub> - clearance of risperidone; V<sub>9-OH-RISP</sub> - Volume of distribution for 9-OH-risperidone; CL<sub>9-OH-RISP</sub> clearance of 9-OH-risperidone; $\beta$ -beta coefficient; $\omega$ - coefficient of variation of inter-individual variability (expressed as a percentage); $\sigma$ - coefficient of variation of residual unexplained variability (expressed as a percentage); *tAge* – age log transformed and centred around the mean; *RSE* relative standard error; *ne* not estimated
\*\*\* p<0.0001

**Supplementary Fig 1.**
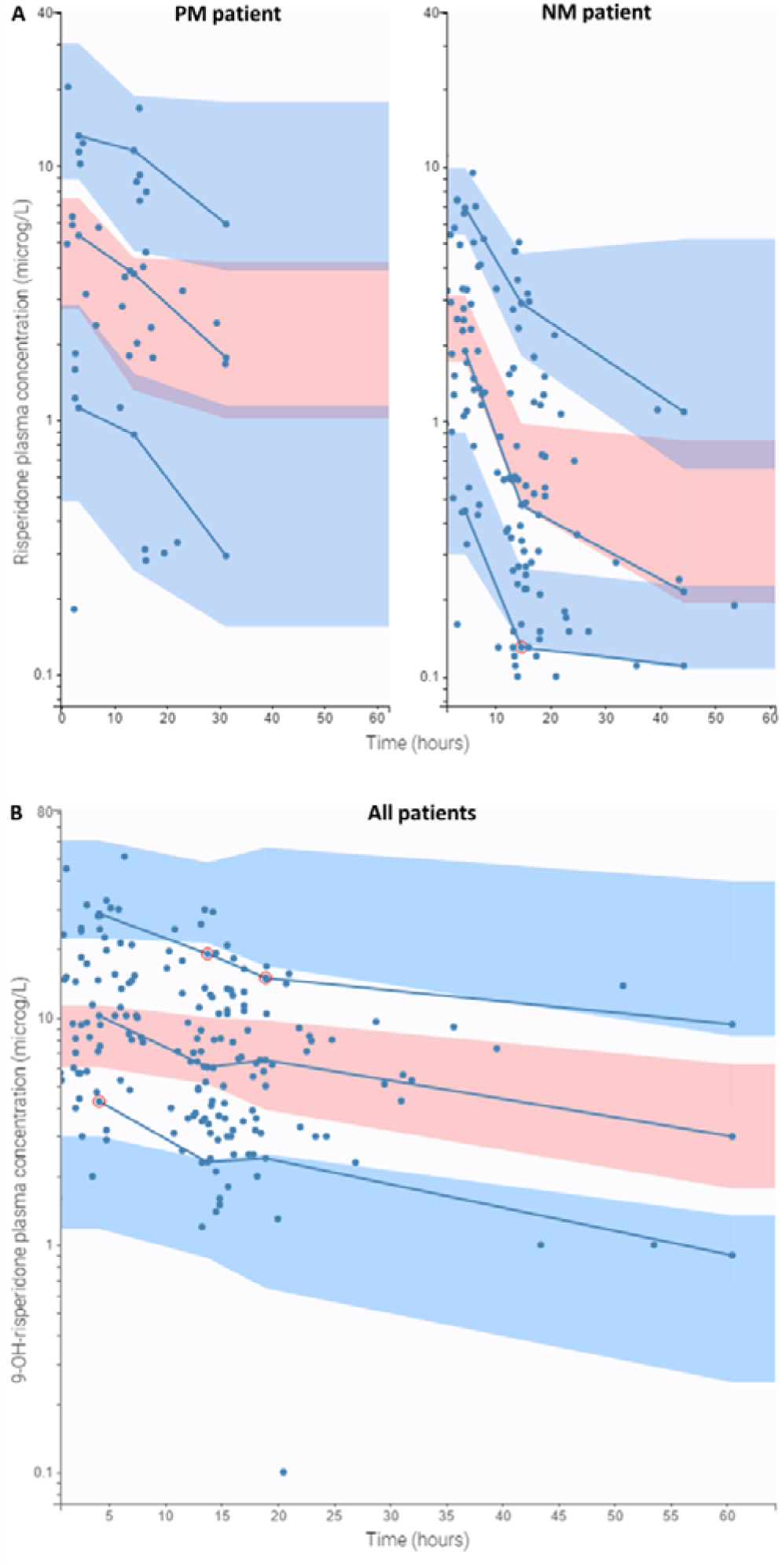
Visual predictive checks. Visual predictive checks (VPC): 95% prediction intervals around the 10th (lower blue shaded area), 50th (pink shaded area) and 90th (upper blue shaded area) percentiles are shown for the final model overlaid to observed data for risperidone in A) Functionally poor metabolisers and B) Functionally normal metabolisers. Each blue circle represents a single plasma sample. Blue lines represent the empirical predictions for each percentile and outliers are circled in red.

**Supplementary Table 2:** Pharmacokinetic Biomarkers and Functional Metaboliser Status.

|  |  |  |  |
| --- | --- | --- | --- |
| **Biomarker** | **Functionally Poor, PM(n=18)** | **Functionally Normal, NM (n=82)** | **Mann Whitney U, p value** |
| **Mean (SD) Peak plasma concentrations, ng/mL** |  |  |  |
| Risperidone | 5.4 (3.2) | 3.6 (2.4) | 0.005 |
| 9-OH-Risperidone | 12.7 (7.6) | 9.5 (8.2) | 0.03 |
| Active Moiety | 17.6 (9.1) | 12.4 (9.9) | 0.007 |
| **Mean (SD) Trough plasma concentrations, ng/mL** |  |  |  |
| Risperidone | 3.9 (2.3) | 1.5 (1.4) | <0.0001 |
| 9-OH-Risperidone | 12.5 (7.5) | 9.2 (8.1) | 0.02 |
| Active Moiety | 16.3 (8.7) | 10.7 (9.3) | 0.003 |
| **Mean (SD) Average plasma concentrations, ng/mL** |  |  |  |
| Risperidone | 2.6 (1.6) | 0.5 (0.8) | <0.0001 |
| 9-OH-Risperidone | 12.1 (7.4) | 8.6 (7.8) | 0.01 |
| Active Moiety | 14.6 (8.0) | 9.0 (8.4) | 0.002 |
| **Mean (SD) trough active moiety concentration: dose (C/D) ratio, ng/mL per mg/day** | 20.2 (7.2) | 7.6 (4.9) | <0.0001 |

## Notes

### Competing Interest Statement

The authors have declared no competing interest.

### Author Declarations

The research was approved by the CATIE protocol and ethics committees, and was reviewed by the National Institute of Mental Health Data Safety and Monitoring Board

## References

1. Ropacki SA, Jeste DV. Epidemiology of and risk factors for psychosis of Alzheimer’s disease: a review of 55 studies published from 1990 to 2003. Am J Psychiatry. 2005;162(11):2022–30.

2. Connors MH, Ames D, Woodward M, Brodaty H. Psychosis and Clinical Outcomes in Alzheimer Disease: A Longitudinal Study. Am J Geriatr Psychiatry. 2018;26(3):304–13.

3. Ohno Y, Kunisawa N, Shimizu S. Antipsychotic Treatment of Behavioral and Psychological Symptoms of Dementia (BPSD): Management of Extrapyramidal Side Effects. Front Pharmacol. 2019;10:1045.

4. Tampi RR, Tampi DJ, Balachandran S, Srinivasan S. Antipsychotic use in dementia: a systematic review of benefits and risks from meta-analyses. Ther Adv Chronic Dis. 2016;7(5):229–45.

5. Ballard C, Howard R. Neuroleptic drugs in dementia: benefits and harm. Nat Rev Neurosci. 2006;7(6):492–500.

6. Schneider LS, Tariot PN, Dagerman KS, Davis SM, Hsiao JK, Ismail MS, et al. Effectiveness of atypical antipsychotic drugs in patients with Alzheimer’s disease. N Engl J Med. 2006;355(15):1525–38.

7. Howard R, Costafreda SG, Karcher K, Coppola D, Berlin JA, Hough D. Baseline characteristics and treatment-emergent risk factors associated with cerebrovascular event and death with risperidone in dementia patients. Br J Psychiatry. 2016;209(5):378–84.

8. Reeves S, McLachlan E, Bertrand J, D’Antonio F, Brownings S, Nair A, et al. Therapeutic window of dopamine D2/3 receptor occupancy to treat psychosis in Alzheimer’s disease. Brain. 2017;140(4):1117–27.

9. Reeves S, Bertrand J, McLachlan E, D’Antonio F, Brownings S, Nair A, et al. A Population Approach to Guide Amisulpride Dose Adjustments in Older Patients With Alzheimer’s Disease. J Clin Psychiatry. 2017;78(7):e844-e51.

10. Katz I, de Deyn PP, Mintzer J, Greenspan A, Zhu Y, Brodaty H. The efficacy and safety of risperidone in the treatment of psychosis of Alzheimer’s disease and mixed dementia: a meta-analysis of 4 placebo-controlled clinical trials. Int J Geriatr Psychiatry. 2007;22(5):475–84.

11. Mauri MC, Paletta S, Di Pace C, Reggiori A, Cirnigliaro G, Valli I, et al. Clinical Pharmacokinetics of Atypical Antipsychotics: An Update. Clin Pharmacokinet. 2018;57(12):1493–528.

12. de Leon J, Wynn G, Sandson NB. The pharmacokinetics of paliperidone versus risperidone. Psychosomatics. 2010;51(1):80–8.

13. Feng Y, Pollock BG, Coley K, Marder S, Miller D, Kirshner M, et al. Population pharmacokinetic analysis for risperidone using highly sparse sampling measurements from the CATIE study. Br J Clin Pharmacol. 2008;66(5):629–39.

14. Hiemke C, Bergemann N, Clement HW, Conca A, Deckert J, Domschke K, et al. Consensus Guidelines for Therapeutic Drug Monitoring in Neuropsychopharmacology: Update 2017. Pharmacopsychiatry. 2018;51(1-02):e1.

15. Vandenberghe F, Guidi M, Choong E, von Gunten A, Conus P, Csajka C, et al. Genetics-Based Population Pharmacokinetics and Pharmacodynamics of Risperidone in a Psychiatric Cohort. Clin Pharmacokinet. 2015;54(12):1259–72.

16. Uchida H, Takeuchi H, Graff-Guerrero A, Suzuki T, Watanabe K, Mamo DC. Dopamine D2 receptor occupancy and clinical effects: a systematic review and pooled analysis. J Clin Psychopharmacol. 2011;31(4):497–502.

17. de Leon J. Personalizing dosing of risperidone, paliperidone and clozapine using therapeutic drug monitoring and pharmacogenetics. Neuropharmacology. 2019:107656.

18. Schneider LS, Tariot PN, Lyketsos CG, Dagerman KS, Davis KL, Davis S, et al. National Institute of Mental Health Clinical Antipsychotic Trials of Intervention Effectiveness (CATIE): Alzheimer disease trial methodology. Am J Geriatr Psychiatry. 2001;9(4):346–60.

19. Simpson GM, Angus JW. A rating scale for extrapyramidal side effects. Acta Psychiatr Scand Suppl. 1970;212:11-9.

20. Barnes TR. A rating scale for drug-induced akathisia. Br J Psychiatry. 1989;154:672-6.

21. Panhard X, Goujard C, Legrand M, Taburet AM, Diquet B, Mentre F, et al. Population pharmacokinetic analysis for nelfinavir and its metabolite M8 in virologically controlled HIV-infected patients on HAART. Br J Clin Pharmacol. 2005;60(4):390–403.

22. Duffull SB, Wright DF, Winter HR. Interpreting population pharmacokinetic-pharmacodynamic analyses - a clinical viewpoint. Br J Clin Pharmacol. 2011;71(6):807–14.

23. McLachlan AJ, Pont LG. Drug metabolism in older people--a key consideration in achieving optimal outcomes with medicines. J Gerontol A Biol Sci Med Sci. 2012;67(2):175–80.

24. Sotaniemi EA, Arranto AJ, Pelkonen O, Pasanen M. Age and cytochrome P450-linked drug metabolism in humans: an analysis of 226 subjects with equal histopathologic conditions. Clin Pharmacol Ther. 1997;61(3):331–9.

25. Uchida H, Takeuchi H, Graff-Guerrero A, Suzuki T, Watanabe K, Mamo DC. Predicting dopamine D(2) receptor occupancy from plasma levels of antipsychotic drugs: a systematic review and pooled analysis. J Clin Psychopharmacol. 2011;31(3):318–25.

26. Wang JS, Ruan Y, Taylor RM, Donovan JL, Markowitz JS, DeVane CL. The brain entry of risperidone and 9-hydroxyrisperidone is greatly limited by P-glycoprotein. Int J Neuropsychopharmacol. 2004;7(4):415–9.

27. van Assema DM, Lubberink M, Bauer M, van der Flier WM, Schuit RC, Windhorst AD, et al. Blood-brain barrier P-glycoprotein function in Alzheimer’s disease. Brain. 2012;135(Pt 1):181-9.

28. Uchida H, Suzuki T, Graff-Guerrero A, Mulsant BH, Pollock BG, Arenovich T, et al. Therapeutic window for striatal dopamine D(2/3) receptor occupancy in older patients with schizophrenia: a pilot PET study. Am J Geriatr Psychiatry. 2014;22(10):1007–16.

29. Graff-Guerrero A, Rajji TK, Mulsant BH, Nakajima S, Caravaggio F, Suzuki T, et al. Evaluation of Antipsychotic Dose Reduction in Late-Life Schizophrenia: A Prospective Dopamine D2/3 Receptor Occupancy Study. JAMA Psychiatry. 2015;72(9):927–34.

30. Aworinde J, Werbeloff N, Lewis G, Livingston G, Sommerlad A. Dementia severity at death: a register-based cohort study. BMC Psychiatry. 2018;18(1):355.

